# Inequality in acute respiratory infection outcomes in the United States: A review of the literature and its implications for public health policy and practice

**DOI:** 10.1101/2020.04.22.20069781

**Authors:** Elizabeth Moran, John Kubale, Grace Noppert, Ryan Malosh, Jon Zelner

**Affiliations:** University of Michigan School of Public Health, Ann Arbor, MI; Carolina Population Center, University of North Carolina, Chapel Hill, NC

## Abstract

Seasonal and pandemic respiratory viruses such as influenza and the novel coronavirus (SARS-COV-2) currently sweeping the globe have often been described as ‘equal opportunity infectors’, implying little socioeconomic disparity in susceptibility. However, early data from the COVID-19 pandemic has underscored that the burden of respiratory viruses actually reflect and magnify existing socioeconomic inequalities. We review the literature on socioeconomic and racial disparities in acute respiratory infection (ARI), as well as ARI-associated hospitalization and mortality. Our goal is to identify key principles of the relationship between socioeconomic inequality and ARI outcomes, as well as highlighting poorly understood areas that need to be addressed by research and policy in the wake of the COVID-19 pandemic. We find that there has been descriptive work in this area, but that there is a distinct lack of cohesive methodology in the literature exploring social determinants and ARI. We propose the fundamental cause theory is a useful framework for guiding future research of disparities in ARI and for the design of interventions to alleviate these disparities.

## Introduction

Acute respiratory infections (ARI) cause substantial morbidity and mortality worldwide^1^, both as the result of seasonal epidemics and pandemics, such as COVID-19. Respiratory viruses such as influenza and respiratory syncytial virus (RSV), have often been described as ‘equal opportunity infectors’, implying little socioeconomic disparity in susceptibility. However, early data from the COVID-19 pandemic has underscored that the burden of respiratory viruses actually reflect and magnify existing socioeconomic inequalities^2^. These trends have been made clear in the United States by the alarming disparities in the toll of severe disease and mortality experienced by African-Americans^3^. At the population level, viral and immune factors are necessary pre-conditions for the emergence and transmission of pathogens causing seasonal and pandemic ARI^4^. At the same time, socioeconomic inequalities are clearly key drivers of exposure, severe disease, and mortality. The rapid pace of transmission and mortality resulting from the COVID-19 pandemic has shone a bright light on these disparities. But they are no less acute in the context of other causes of seasonal and pandemic ARI, despite attracting less attention. In this paper, we review the literature on socioeconomic and racial disparities in ARI infection, hospitalization, and mortality in the United States. By focusing on the existing literature on influenza, RSV, and all-cause ARI we hope to identify key principles of the relationship between socioeconomic inequality and ARI outcomes, as well as highlighting poorly understood areas that need to be addressed by research and policy in the wake of the COVID-19 pandemic.

### What are the causes of ARI disparities

Disparities in ARI outcomes may result from 1) differential rates of exposure owing to occupation, housing, and other factors, 2) differential susceptibility to infection upon exposure, owing to both social and medical factors, e.g. access to vaccination where it is available, and 3) differential ARI-associated morbidity and mortality resulting from comorbidities increasing the likelihood of severe disease, limited access to quality care and therapies (e.g. antivirals) once infected. ARI-related disparities falling along these dimensions are evident at the within-country level, and at the global scale: A disproportionate amount of severe ARI-related disease and death occurs in low- and middle-income countries (LMICs), with as much as 99% of in-hospital deaths associated with RSV occurring in LMICs as of 2015^5^. In addition, a 15-fold difference in influenza-associated case-fatality has been observed between LMICs and wealthy countries^6^, underscoring how global inequality exacerbates the toll of death and disability attributable to ARI.

However, a persistent focus on biological dimension of infectiousness has reinforced the perception that that social inequality has little to do with the burden of ARI both domestically and globally. This perception dates back to the pioneering work of McKeown and Record, who ruled out standard of living disparities in ARI mortality in 19^th^-century England and Wales because of their highly-infectious and seasonal nature. As a result, the focus of research on the social determinants of infectious disease has been on inequity in chronic and endemic infections, such as Tuberculosis, diarrheal disease^7^, and HIV. Recently, however, attention has returned to social determinants of ARI, reflected in an increasing number of studies of disparities by race/ethnicity and SES in infection risk, severity and mortality for influenza^8–12^, RSV^13^ and all-cause ARI^14–18^. Nevertheless, our understanding of the mechanistic links between SES and ARI-related risks remains underdeveloped, as does our ability to address inequalities in ARI outcomes.

Closing this gap is urgent, as the question of how to address these disparate outcomes requires us to first understand how they are generated. Our goal is to identify the key axes of SES-related disparity in ARI infection risk and outcomes. In many ways, the urgency of addressing these disparities results from positive momentum in biomedical research against influenza, RSV and other ARIs. A push towards a more broadly protective influenza vaccine, increased use of antivirals to improve outcomes, and advances in genomics and mathematical modeling to understand transmission, can lead to significant strides against influenza and other endemic and emerging viral ARIs. However, such advances in *population-level* protection are often accompanied by a widening of disparities in outcomes, as new preventive measures and therapies become available first to the well-off. As recent discussions of the ability to practice social distancing, and whether a COVID-19 vaccine will become broadly available shows, these issues remain woefully unaddressed, and have significant implications for public health.

## Data and Methods

### Literature Search

We searched PubMed for articles documenting socioeconomic disparities in ARI infection/disease risk, severe outcomes, and mortality. First, we attempted to capture articles investigating disparities in all-cause ARI, influenza-like illness (ILI), and RSV. Then, we identified articles documenting disparities in influenza infection risk and outcomes. We separated these searches to ensure each was sensitive to key differences in the literature, most notably around access to and uptake of influenza vaccination.

We used the PubMed special query for health disparities in conjunction with search terms relating to all-cause ARI, RSV, and influenza (Figure 1). The health disparities query identifies articles evaluating disparities in health outcomes and healthcare access with inequities in dimensions of race/ethnicity, SES, gender identity and sexual orientation, insurance status, and other populations described as “vulnerable.” We then screened studies using title and abstract review to identify articles with a specific focus on all-cause ARI, ILI, RSV, and influenza. Additional articles and reviews were added to our results upon recommendation from topic-area experts. Articles were not excluded based on year of publication. Full search queries are available in the supplementary materials.

**Figure 1:**
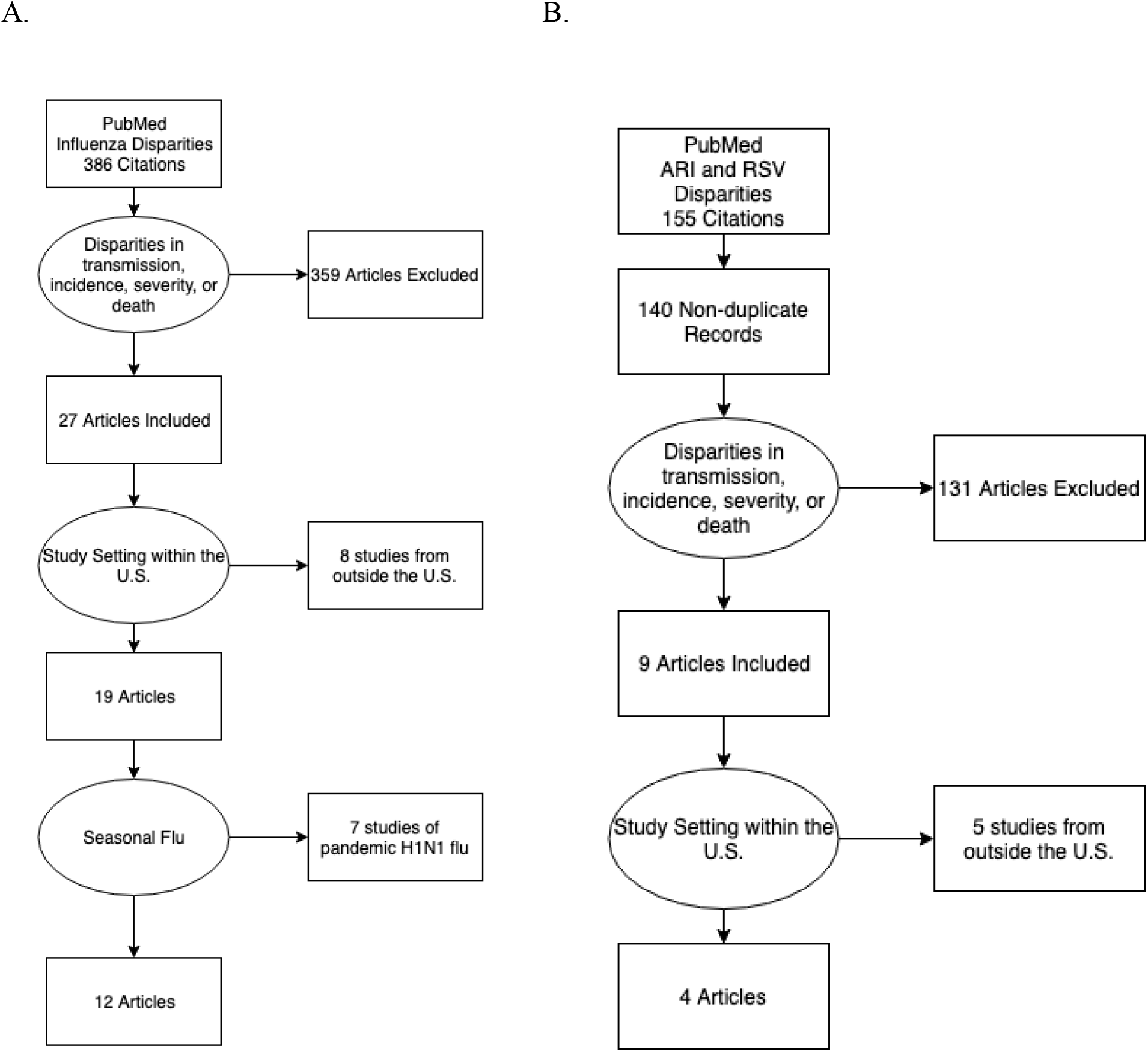
Search results and screening for A. influenza disparities and B. ARI and RSV disparities.

## Results

Our all-cause ARI and RSV search returned 155 papers, of which 9 studies met the inclusion criteria. Of these, only 4 of the retained articles used data from the US. The influenza search returned 386 results, of which 27 were deemed relevant following title and abstract review. Of these, 12 addressed seasonal influenza in the US, with the remaining 15 focused on the 2009 H1N1 influenza pandemic. All selected articles, organized by etiology, dimension of disparity assessed, and measured outcome (incidence, severity, death, etc.) are listed in Table 1.

**Table 1:**
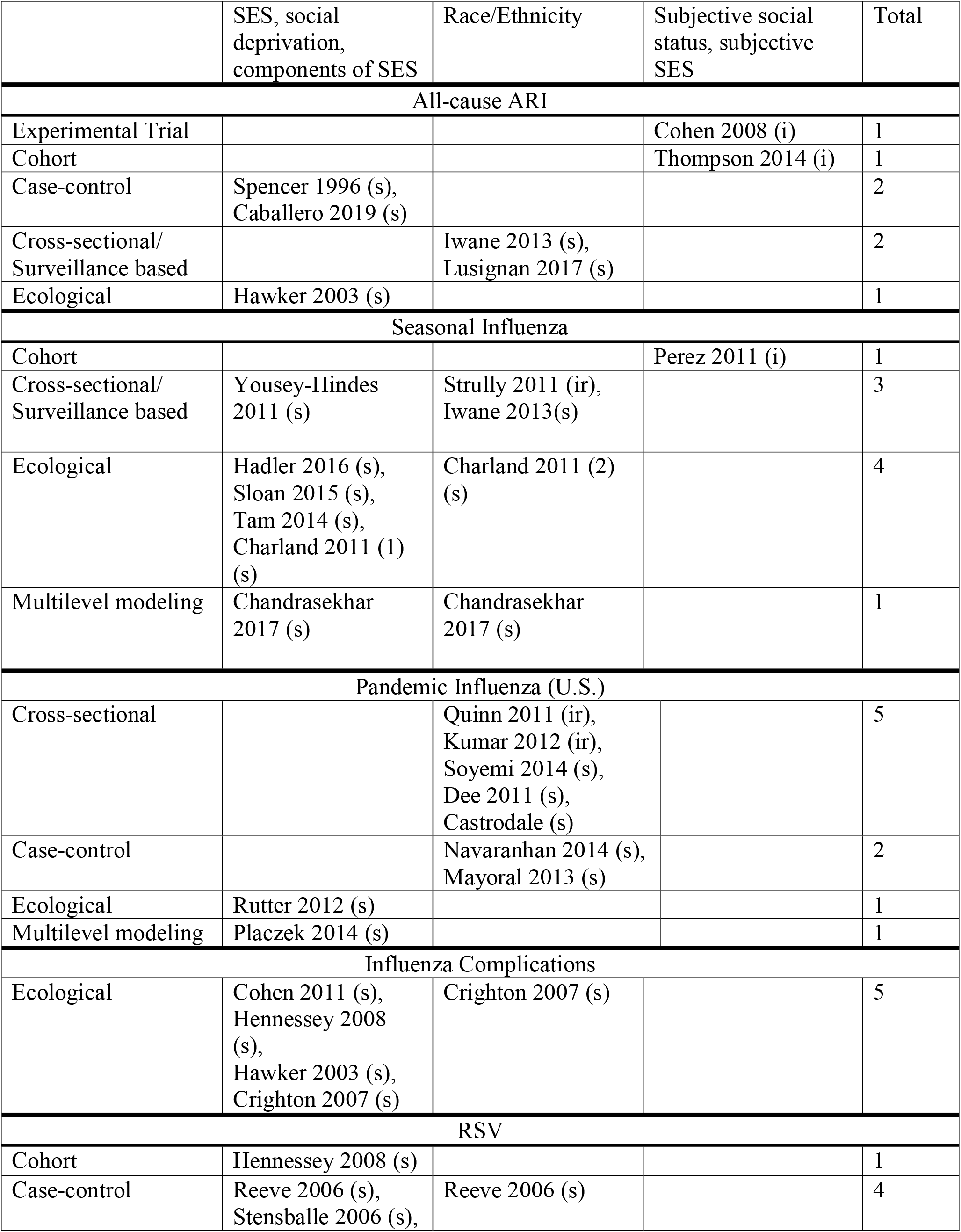
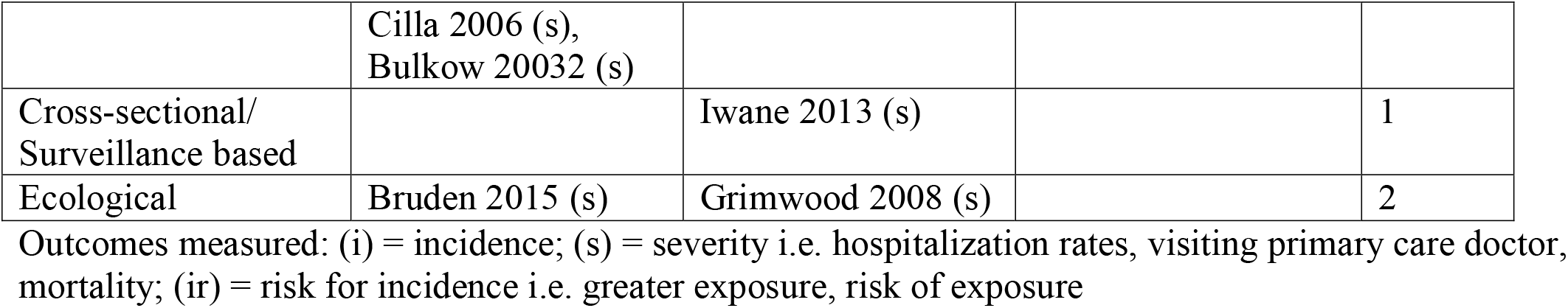
Study design, dimension of disparity, and outcome of articles included.

### All-Cause ARI

A wide range of SES-related disparities in all-cause ARI were identified in studies from the US^14,15,19^, UK^16,17,20^, and Argentina^18^. Two studies reported increased ARI incidence^14,15^ among those with lower self-reported social status^14^ and SES^15^. Race/ethnicity was also found to be associated with increased severity, as indicated by two studies reporting increased ARI-related hospitalization in a US study of children under 5 using data from New Vaccine Surveillance Network (NVSN)^19^ and primary care seeking in UK using sentinel surveillance data^20^. SES disparities were also evident in all-cause ARI-related complications: two studies reported increased hospital admission for ARI^17^ and bronchiolitis^16^ and increased rates of medical intervention due to bronchiolitis^16^ with increasing Townsend deprivation score, index used to quantify material deprivation based on car and house ownership, household overcrowding, and employment, with a higher score showing more material deprivation^16,17^. Finally, one study reported increased under-5 mortality due to ARI among those with proxy measures for poverty including having an adolescent mother, household crowding, and lacking running water in home^18^.

### Incidence

In a multi-state cohort study, female healthcare workers from Texas, Washington, and Oregon, with lower self-reported social status reported higher rates of ARI symptoms ^14^. In an experimental study, higher subjective SES was protective against the development of cold symptoms after exposure to rhinovirus or influenza virus^15^.

### Severity

Disparities in severity of ARI were indicated using hospitalization and care seeking rates in a variety of study designs. Using population-based surveillance data, the NVSN study found increased hospitalization rates for all-cause ARI among Black children as compared to white children; however, these results were not adjusted for SES^19^. Additional surveillance data from the Royal College of General Practitioners Research and Surveillance Centre in England found racial and ethnic minorities were more likely to seek care for cold/flu symptoms than whites^20^. In the UK, a case-control study matching children under 1 year old admitted to Sheffield hospital with clinical suspicion of bronchiolitis to healthy community controls identified from birth records found that children living in the two most deprived electoral wards, as defined using the Townsend index, were 1.5 times as likely to be admitted to the hospital and 1.7 times as likely to require a medical intervention than children living in other parts of the city^16^. An ecological study in the West Midlands Region of the UK found neighborhoods with higher Townsend scores of deprivation experienced higher rates of childhood hospital admissions due to ARI and pneumonia^17^.

### Mortality

A case-control study in Buenos Aires found death due to ARI in children under 5 was associated with living in a crowded household, having an adolescent mother, and lacking running water in the home^18^. No other papers discussed mortality due to all-cause ARI.

### Seasonal Influenza

Studies of disparities in seasonal influenza outcomes have focused primarily on the relationship between neighborhood SES and rates of hospitalization for severe disease in^8–12^ with fewer studies of individual-level outcomes^21^. In addition, studies have investigated inequalities in influenza-related healthcare utilization (e.g. visits to outpatient clinics)^21,22^, incidence^23^, and exposure^24^ as they relate to SES, race/ethnicity^9,19,22^, and socioemotional stress^23^. SES and exposure indicators utilized in these studies include percentage of households living in poverty^8,10–12^, female headed households^9,10^, household crowding^9,11^, and neighborhood population density^10^.

### Incidence

Two papers described disparities in rates of influenza susceptibility and exposure in older African Americans^24^ and college students with higher perceived stress^23^. Black Americans in nursing homes were less likely to be vaccinated against seasonal influenza, and were less likely to have vaccinated contacts, due to racial and socioeconomic segregation in nursing home care. The authors of this study argue that the results from nursing homes may provide clues to residential segregation’s effects on seasonal influenza risk at the population level^24^. In a cohort study of U.S. college students, an increase in perceived stress score was associated with a 25% greater rate of self-reported ILI, after adjustment for demographic factors, behaviors and flu vaccination^23^.

### Severity

Hospitalization was the most frequently studied dimension of severity used to characterize influenza disparities, followed by overall healthcare utilization^21,22^. Area-level poverty and race/ethnicity were used as predictors of risk in a number of studies examining disparities in rates of influenza-related hospitalization. Two ecological studies found that living in neighborhoods with a high percentage of residents below the poverty line and in female-headed households experienced higher rates of influenza hospitalization ^8,9^. Similarly, census-tract rates of influenza-associated hospitalization surrounding Nashville, Tennessee, increased with population density and the percentage living in poverty, female headed/crowded households ^10^. In New Haven, Connecticut, the annual incidence of pediatric influenza-associated hospitalizations in high-crowding census tracts (defined as >5% of households with > one occupant per bedroom), and high-poverty census tracts (defined as >20% of households below the poverty line), was > 3 times higher than in low-crowding/low-poverty census tracts^11,12^. This disparity remained even after adjustment for comorbid medical conditions and influenza vaccination^11,12^. In a study of individual-level outcomes using data from FluServ-NET, across multiple US counties and influenza seasons, African American and Latino adults had increased odds of influenza-related hospitalization than whites^9^. One study using population-based surveillance data found significantly higher hospitalization rates for laboratory-confirmed influenza for Black children than for white children^19^.

### Pneumonia

Viral and bacterial pneumonias are common ARI-related complications that reflect SES-based disparities in baseline health status as well as access to care. Several studies identified by our search focused specifically on disparities in hospitalization^13,17,25,26^ and mortality^27^ associated with diagnosis of pneumonia and influenza (P&I).

### Severity

In a study examining socio-demographic factors increasing risk of P&I hospitalization in elderly populations using Medicare data from 1991-2004, counties with high proportions of live-in grandparent caregivers and lower median income experienced higher rates of pneumonia-related hospitalization^26^. In a study of Alaska Natives using hospitalization records from 2000-2004, regions with a lower proportion of households with in-home water service, indicating higher poverty rates, also had higher hospitalization rates for P&I^13^.

### Mortality

There is some evidence that disparities in P&I associated mortality have decreased over time. One study using age-adjusted mortality rates for P&I show a decreasing ratio and absolute difference between Black and white Americans from 1950 to 2000^27^. The authors attribute this reduction in disparity due to widely available treatment to guard against P&I complications and mortality^27^.

### RSV

Literature examining RSV-specific disparities was limited to hospitalization/severity. Disparities were identified in dimensions of race/ethnicity^19,28,29^, household socioeconomic status^13,30^, high population density^30^, maternal unemployment^31^, younger maternal age^32^, and household crowding^33^. A study using population-based surveillance data found that hospitalization rates for laboratory confirmed RSV were similar for Black and white children under 12 months of age, but for children > 12 months, hospitalization rates for lab-confirmed RSV were higher for Black children than for white children^19^. A study utilizing surveillance and hospital admittance data in Southwestern Alaska identified villages with lower proportion of houses plumbed water, higher proportion of household crowding, and higher proportion of families living below the poverty line as risk factors for hospital admission for RSV^30^. Similar risk factors were also identified in non-US studies^29,28,31 32^.

### Pandemic Influenza

Pandemics of influenza and other viral respiratory pathogens, such as COVID-19, are characterized by higher overall attack rates, elevated morbidity and mortality, unevenly adopted countermeasures, and increased strain on health systems. All of which may magnify inequalities in patterns of disease, despite a complete or near-complete lack of protective immunity. Because of this lack of acquired immunity in the population, the age distribution of infection is likely to be different than seasonal epidemics, resulting in differential patterns of healthcare utilization and mortality risk. Nevertheless, in a pattern echoing findings for seasonal flu, RSV and all-cause ARI, inequalities in the most severe ARI pandemic outcomes have consistently been documented. For example, historical data from Chicago illustrated disparities in 1918 influenza pandemic mortality by neighborhood SES and racial composition^34,35^, with some of this disparity explained by differential rates of transmission in low-SES, overcrowded neighborhoods^34^. In 2009, a number of studies documented social race/ethnic outcome disparities^36–41^, with lower SES^41^ associated with increased exposure risk^36^, as well as overall incidence^37^, hospitalization^38,39,41^, complications^36^, and death^39,40^ due to pandemic H1N1.

### Incidence

A nationally representative survey from the U.S. measured risk factors for exposure during the 2009 pandemic, including living in an apartment building, relying on public transportation, and difficulty finding day care that was separate from other children. This study found that these risks were significantly more common among non-white participants^36^. Another US study identified a lack of access to sick-leave and increased number of children in the household as risks for higher ILI incidence during the H1N1 pandemic^37^. The authors determined that this increased risk was associated with Hispanic ethnicity even after controlling for education and income^37^, suggesting that additional, unmeasured SES-related risk factors contributed to this disparity.

### Severity

In a study using surveillance data from Illinois, hospitalization rates were twice as high for Hispanics and Blacks compared to whites during the 2009 H1N1 pandemic^38^. An analysis using data from multiple nationally representative surveillance networks investigated disparities in hospitalization and death found substantial differences in morbidity and mortality between race/ethnic groups^39^. Age-adjusted hospitalization rates for minorities were two times higher than those for non-Hispanic whites.

### Mortality

One study using nationally representative surveillance networks found non-Hispanic Black and Hispanic children had disproportionately higher mortality than non-Hispanic whites^39^. During the H1N1 pandemic, American Indians and Alaskan Natives had a mortality rate 4 times higher than all other racial/ethnic groups^40^. Conversely, a study using hospital discharge and census data found the living in a higher poverty census tract (>6% below poverty line) and Hispanic ethnicity was associated with lower risk of ICU admission for H1N1 related hospitalizations in Massachusetts^41^, although it is unclear if this result reflects differential severity or disparities in access to care.

### Outside the United States

Research outside of the United States has identified similar relationships between inequality and pandemic ARI outcomes. For example, in a case-control study in Ontario, Canada of persons hospitalized for ARI during the 2009 H1N1 pandemic, the odds of H1N1 infection were higher among adults identifying as East/Southeast Asian, South Asian, and Black compared to whites^42^. In Spain, individuals belonging to an ethnic minority group were more than twice as likely than whites to be admitted to the hospital, and individuals with a secondary or higher education were 0.54 times less likely to be admitted to the hospital as individuals with less education^43^. In England, the 2009 H1N1 pandemic caused 3.1 and 2.0 times higher mortality rates in the two most socio-economic deprived quintiles compared to the least deprived quintile, and this disparity persisted after adjustment for underlying medical conditions and age^44^.

## Discussion

The COVID-19 pandemic has shown in stark relief the ways in which SES and racism structure ARI outcomes at every level from exposure to disease severity and mortality. However, despite the large toll of mortality exacted by seasonal and pandemic ARI and the clear evidence of disparities, the number of studies investigating disparities in ARI outcomes and incidence is too limited to help chart a way towards more equitable outcomes. Notably, most of the analyses discussed in this review come from data collected by studies in which identifying disparities was not a primary goal, and thus was not explicitly incorporated into study design and the enrollment of participants. Further, much of the previous work has documented disparities, for example by race/ethnicity, but has not explored the history of policies and practices that specifically marginalized these racial/ethnic minorities and led to disparate outcomes^45^. This echoes earlier findings that studies focusing on identifying and addressing social determinants are underrepresented relative to the importance of social factors in structuring infectious disease risk^46^.

The lack of studies dedicated to identifying and ameliorating ARI disparities guarantees that the analyses reviewed here provide a decidedly incomplete picture of the drivers of disparities in ARI-related outcomes. For example, many of the studies identified in our search is that they characterized disparities among the population of cases presenting at a point of care, ranging from routine medical visits to hospitalization. This reliance on clinical data is likely to under-count risk in populations without access to care, who are disproportionately likely to have low SES, and/or come from marginalized groups, e.g. undocumented immigrants, for whom real and perceived overlaps between medical and legal systems may serve as a disincentive to care-seeking. Finally, most ARI cohort studies, which allow for granular examination of individual-level risk and protective factors, drew largely on geographically concentrated study populations that are largely homogeneous in terms of SES and race/ethnicity.

### A conceptual framework for unraveling ARI disparities

It is evident from our review that the lack of a coherent theoretical framework for understanding relationships between social inequality and ARI incidence, severity, and mortality is a significant factor in the dearth of research in this area. The evidence turned up by our review, as well as the issues of racially and socioeconomic disparate exposure risks, coupled with unequal access to testing and treatment made clear by the COVID-19 pandemic, demonstrate that the principles underlying Link & Phelan’s theory of SES and race as fundamental causes of health and illness apply to ARIs as much as many non-communicable diseases as well as infections more classically understood has having social antecedents, such as tuberculosis and diarrheal disease. Fundamental cause theory (FCT) posits SES drives disparities through its impact on the financial and social power to marshal the material and social resources that are protective of health^47^. FCT is a mainstay of the literature on SES-related disparities in non-communicable diseases, but has enjoyed very limited application to infectious diseases^48^. A key tenet of FCT is that these disparities are persistent and are likely increase even as population-level risk goes down. For example, medical innovation is likely to increase disparities because new interventions typically become available first to the well-off and well-insured. This strongly suggests that progress in the treatment and prevention of ARIs using antivirals, monoclonal antibodies^49^, influenza vaccination, and molecular genotyping to inform surveillance and control, are likely to result in increased inequality in both ARI incidence and severe outcomes. These issues have once again been front-and-center during the COVID-19 pandemic, as well-off individuals in the United States enjoyed preferential access to testing in the early days of the pandemic. Similarly, the ability to participate in ‘social distancing’ by working from home, having food and other essential delivered to one’s home, and engage other protective measures are clearly a function of power and material resources. In the absence of effective vaccines and antivirals, this is the key technology available to protect oneself from an emerging infection. As preventive and therapeutic interventions against COVID-19 become available, we are likely to see these also distributed preferentially to those with greater means, thereby only exacerbating the yawning disparities in exposure, severity, and mortality that are already evident.

### Implications for Study Design

Surveillance and large network studies can estimate burden of disease and influenza vaccine effectiveness (VE), but may under enroll low-SES and non-white populations and are by design limited to those with access to care since participants are enrolled at the point of care. Ideally, large longitudinal cohort studies with study samples that are representative of the US population would be available to estimate differences in incidence and risk and VE. A meta-analysis found that 97% of observational studies on influenza severity were cohort studies, but very few of these studies collected information on race/ethnicity and the authors do not report on measures of SES in their analysis^55^. To capture the components of risk associated with social disparities, these studies would ideally span multiple seasons and account for known factors associated with risk such as age, influenza infection and vaccination history, household composition, crowding, and other potential exposure and susceptibility related factors. In the absence of such studies, greater attention is needed to increase the socioeconomic diversity of the populations that studies of seasonal ARI risk and VE draw upon, as well as the potential impact of SES on infection outcomes and vaccine effectiveness. Where such data are not available, careful statistical modeling is essential. For example, approaches such as multiple regression with poststratification (MRP), which can be used to make unbiased population-level predictions from non-representative data, may help address some of these questions while we wait for more detailed data^56^.

### Implications for Action

FCT highlights why identifying and intervening only on mediators of the relationship between SES and disease is unlikely to be effective: risk of ARI infection, severe disease, and mortality cannot be reduced to the impact of a single variable, e.g. vaccination. Instead, it reflects a spectrum beginning with comorbid health conditions, the ability to avoid exposure, access to preventive and therapeutic treatment, and other factors. These issues are arguably even more acute for infectious diseases than for non-communicable ones, as the risks of our friends and family directly impact our own risks via transmission.

An FCT perspective suggests that effective policies must address inequality in access to prevention and treatment, as well as inequity in the ability to avoid exposure. For example, policies limiting or eliminating paid sick leave have been shown to increase rates of influenza transmission in the workplace, and to disproportionately affect racial/ethnic minorities^37^, with similar patterns evident in the COVID-19 pandemic. In order to be able to reduce transmission, symptomatic persons should be treated with antivirals (when applicable) and practice home isolation^57^. This cannot be achieved without policies that support rather than penalize such behavior. The lack of supportive workplace policies has been suggested to result in a population-attributable risk of 5 million additional cases of ILI in the US with a disproportionate burden on Hispanic Americans^37^. The lack of a health system ensuring equal access to prevention and care is clear a key factor in exacerbating SES-related disparities in seasonal ARI incidence, as well as undermining pandemic preparedness. However, in the absence of sweeping changes that would address the root causes of these disparities, targeted vaccination for influenza^58,59^ and RSV^60^ based on age specific contact and transmission patterns has been theorized to improve both the efficacy of vaccines. Identifying high priority groups for vaccination has been shown to be effective in reducing disease burden when target groups are those with high-risk of infection or high transmission potential for both RSV and influenza^58–61^. The expansion of targeted programs to improve access to vaccination among those of lower SES could be beneficial in reducing disparities. When identifying groups for targeted vaccination, especially those of disadvantaged backgrounds, it is important to address past abuses that may lead to mistrust of these programs. Finding ways to engage with organizations that serve low SES and racial minority communities to address concern and reduce vaccine hesitancy will be paramount to the success of these programs^36^. New vaccine rollouts, such as an RSV vaccine and a universal flu vaccine, should include some targeted vaccination programs to ensure populations already at increased risk of infection are given access to new technologies to prevent the intensification of disparities. Nevertheless, FCT suggests that the long-term efficacy of such interventions is likely to be limited if the upstream drivers of inequality are not addressed: New pathogens and technologies will inevitably expose the weaknesses of existing strategies, once again opening up a gulf between the well-off and everyone else.

### Conclusion

In sum, our findings highlight wide disparities in ARI outcomes by socioeconomic status and race/ethnicity in the U.S. Though the COVID-19 pandemic has brought many of these concerns to the forefront of the public and scientific consciousness, it has also shown how poorly developed our set of tools for addressing both the root and proximal causes of these disparities is. This study represents a first step towards the development a coherent framework for identifying disparities in ARI and their causal antecedents. This will hopefully increase clarity around when and where socially focused interventions (i.e. healthcare and workplace policies) should be prioritized, as well as how biomedical innovations (e.g. vaccination, antivirals) should be distributed to shrink group-level disparities at the same time that they minimize population risk.

To accomplish this, a sustained effort to build studies and create standardized measures purposively aimed at understanding and curbing SES-related disparities in ARIs is urgently needed. However, even with this lack of consensus, the body of evidence suggests there are disparities in ARI incidence, severity, and mortality. With potential new technologies on the horizon to prevent and better treat ARIs from RSV to influenza as well as emerging infections with pandemic potential, such as COVID-19, the time to make a plan for how to ensure the equitable distribution of benefits from these and other technologies is now.

**Table 2:**
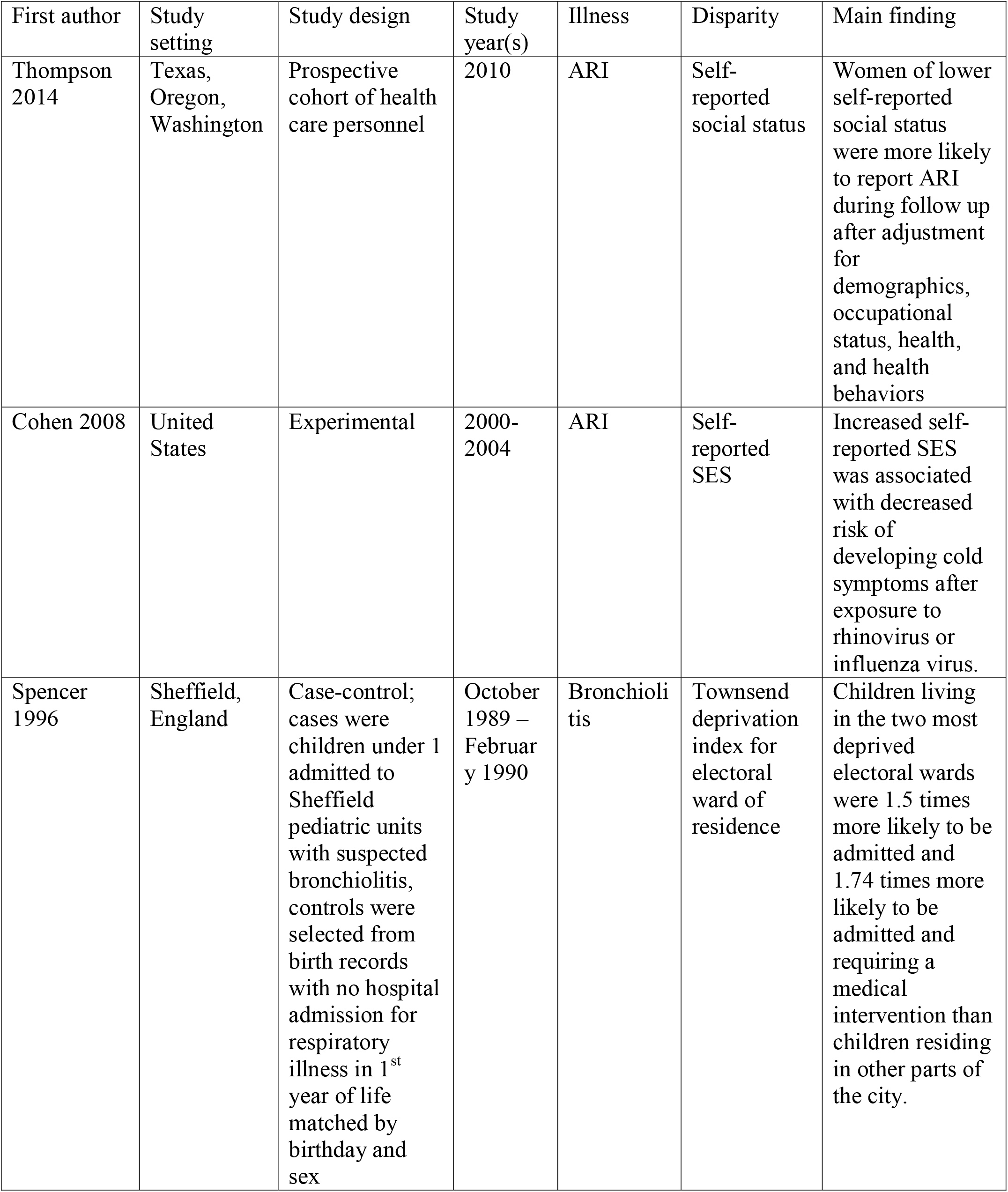
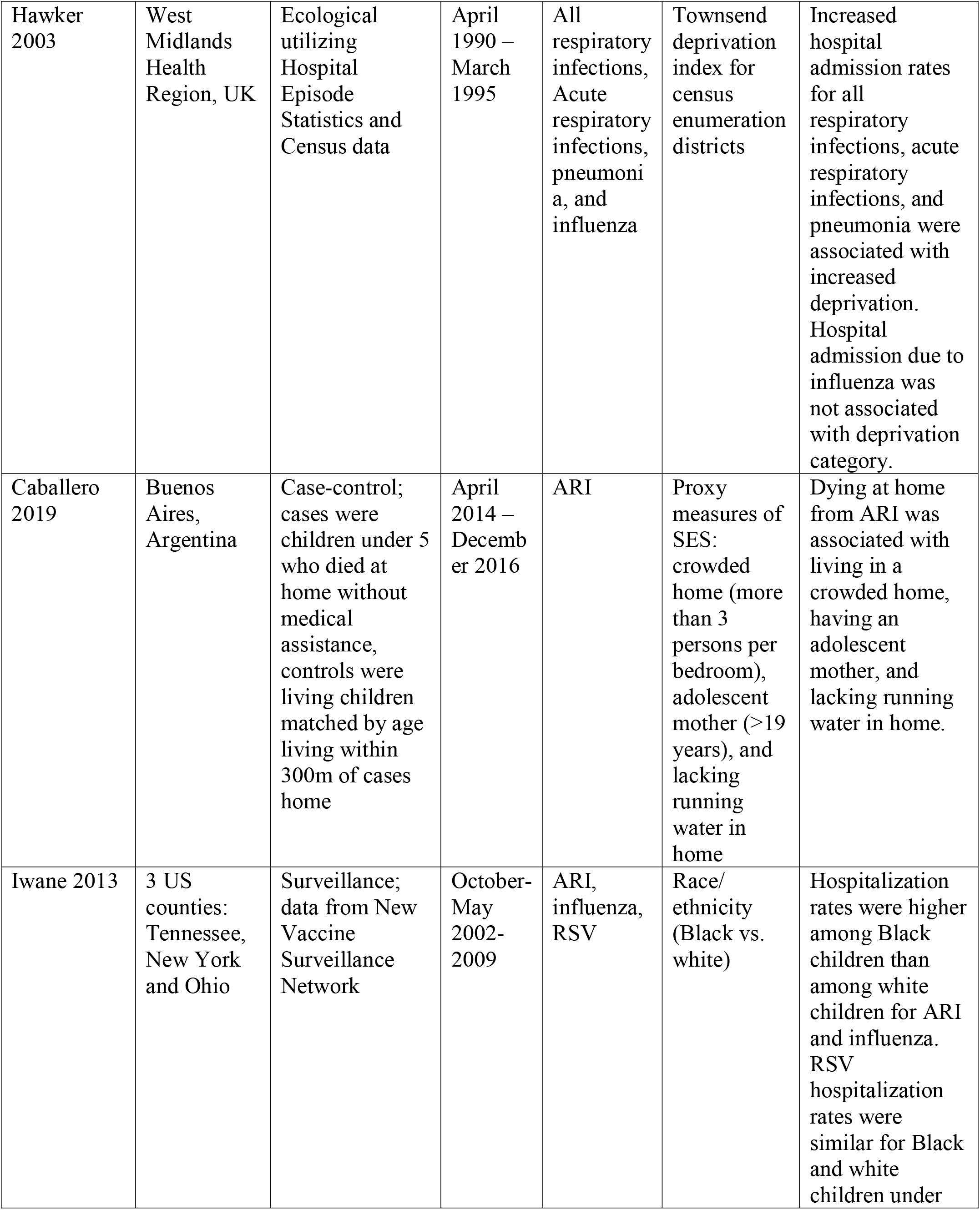
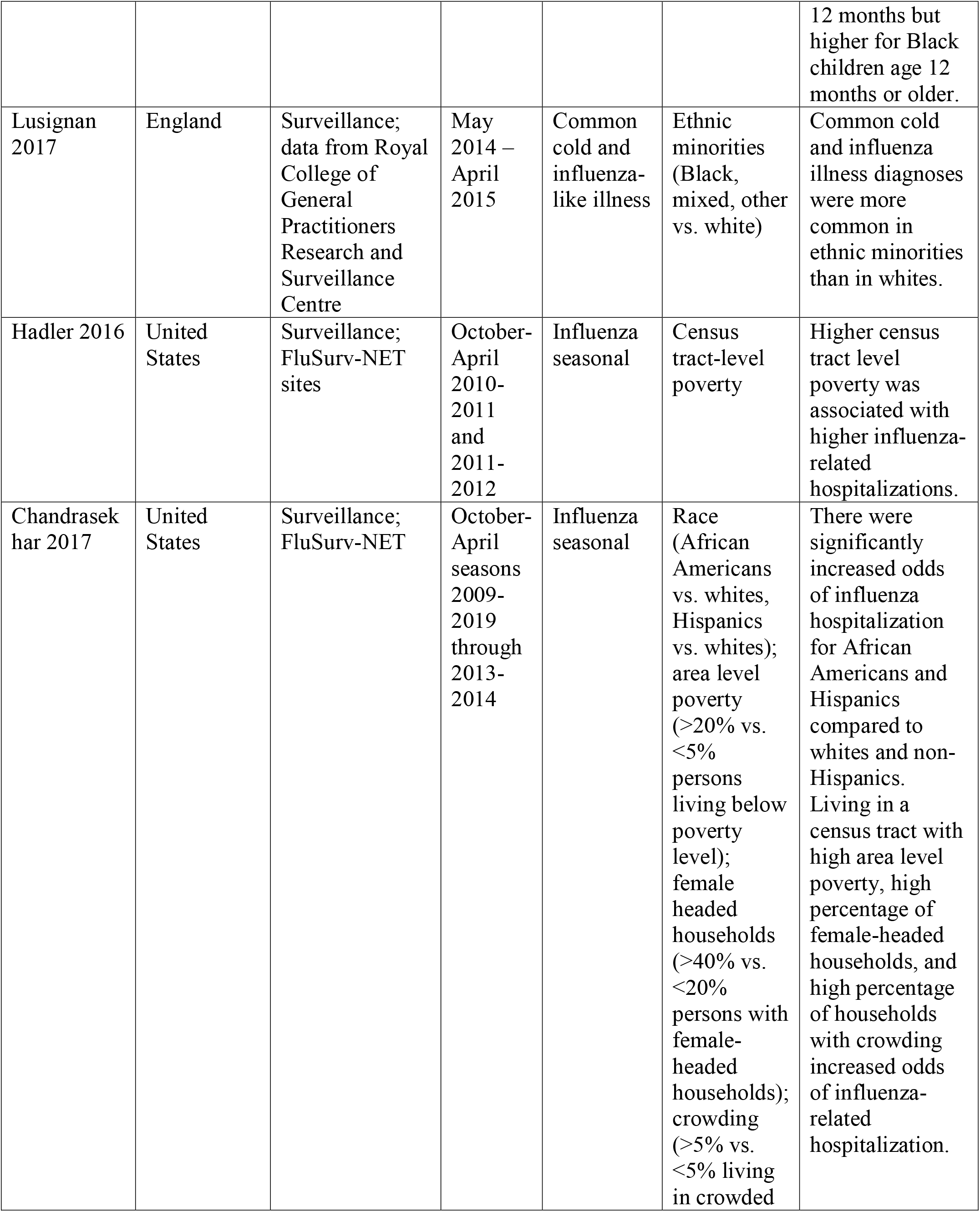
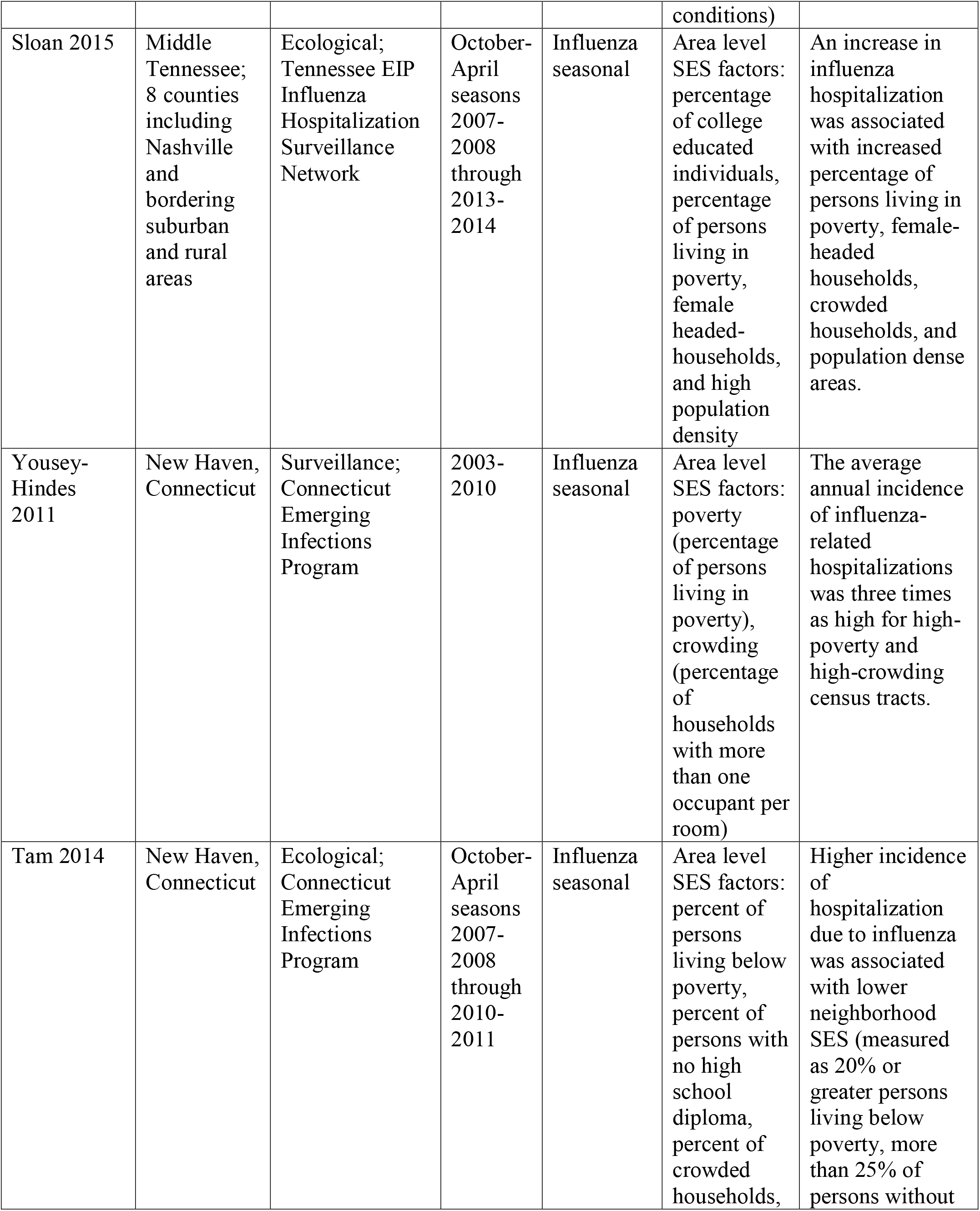
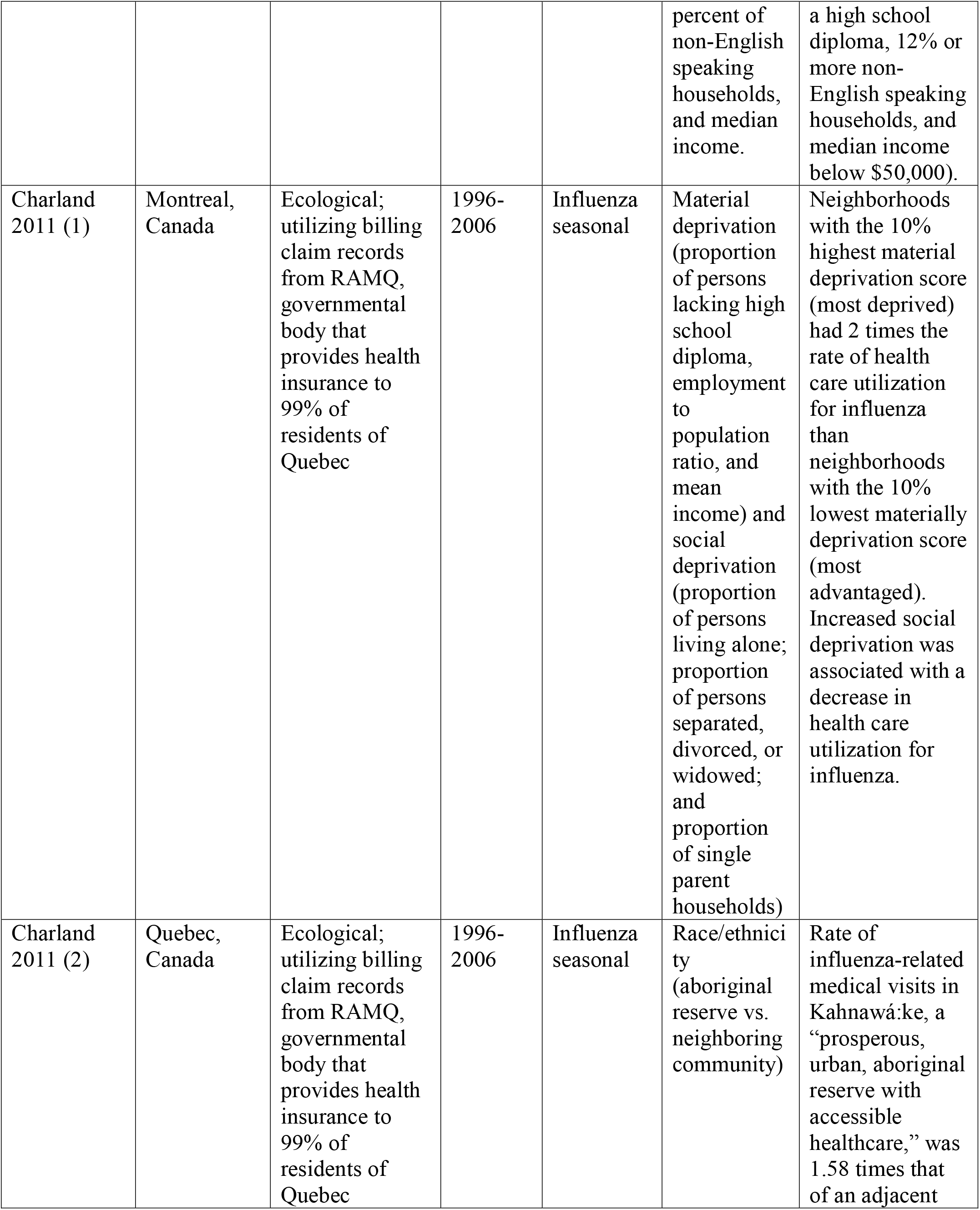
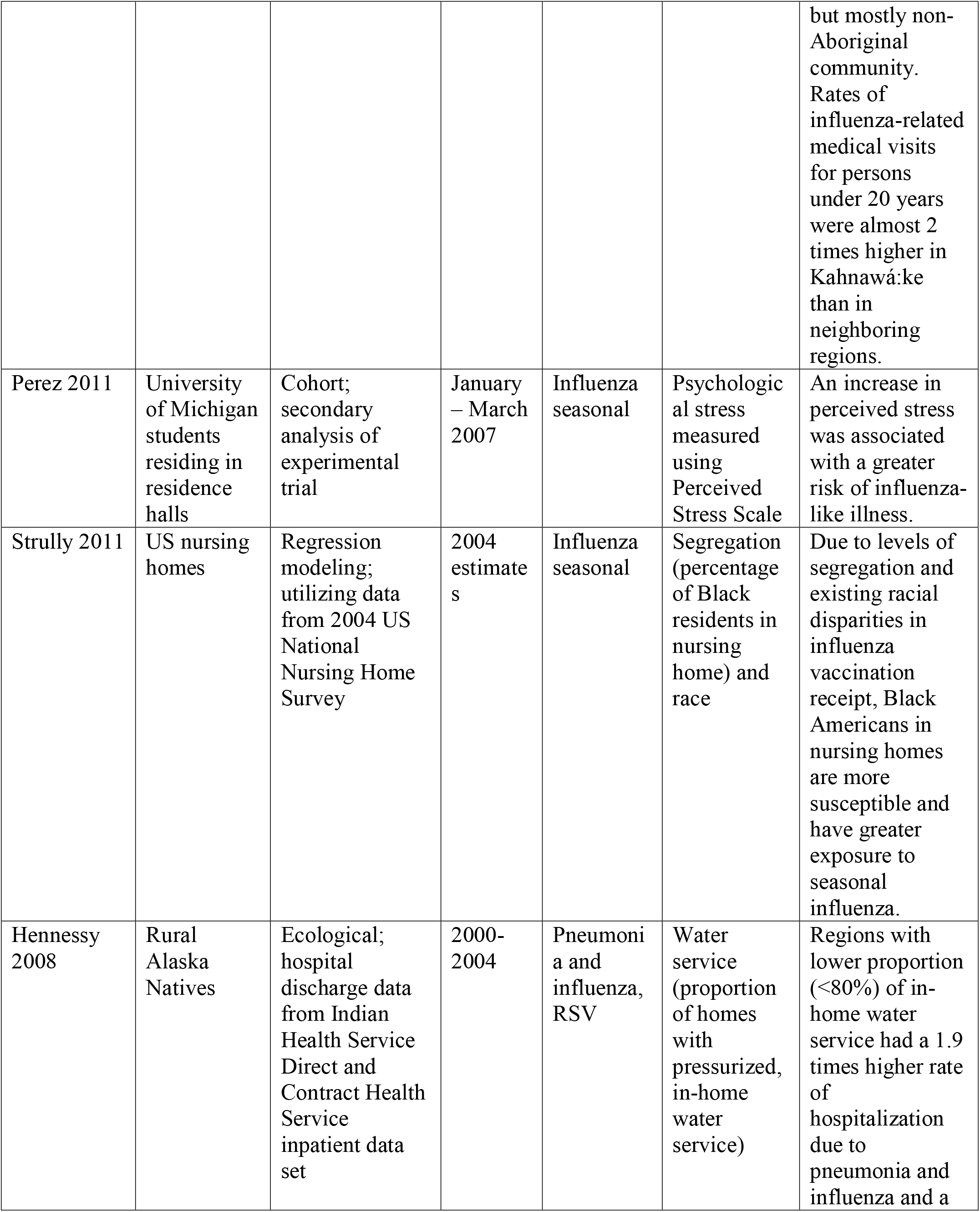
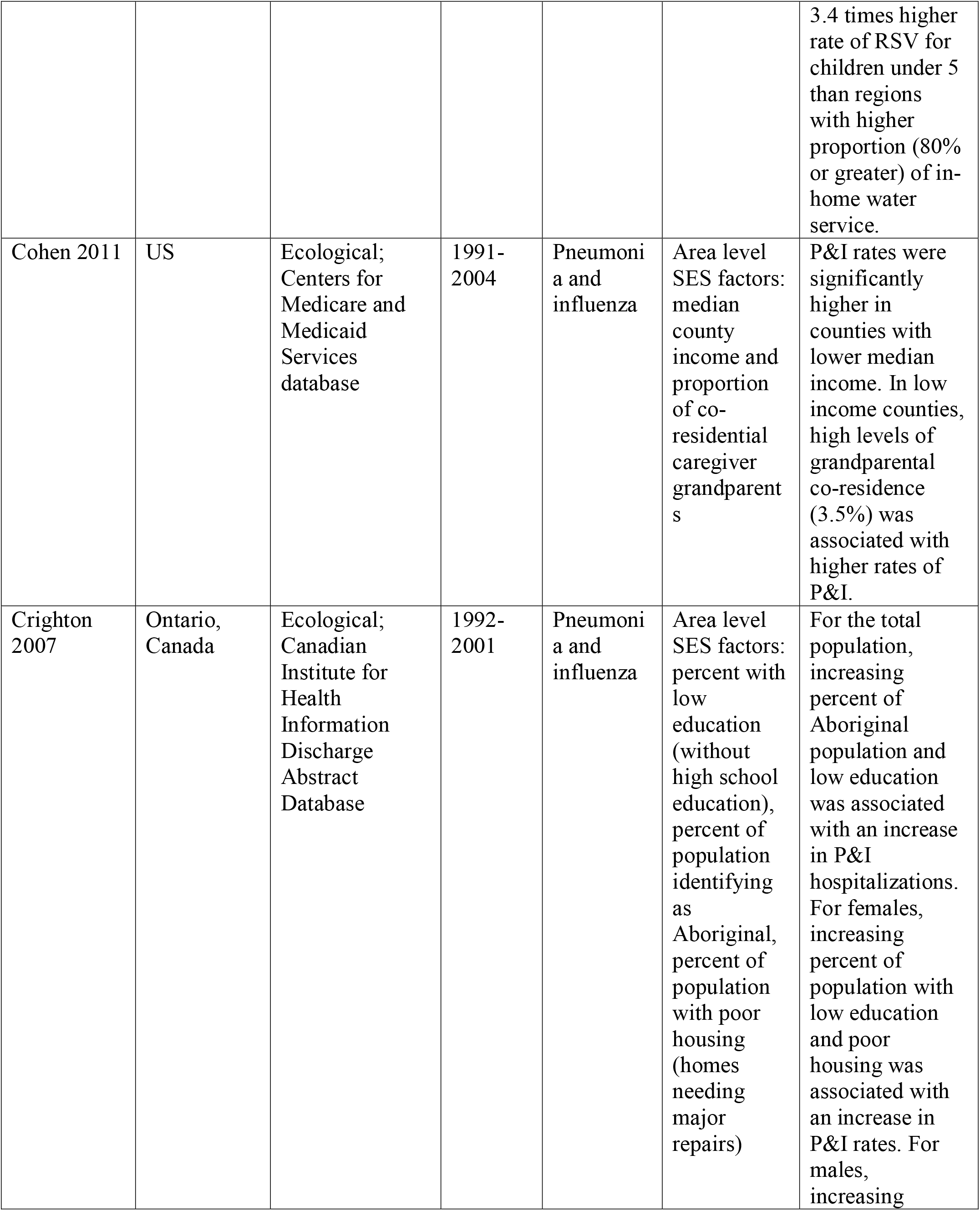
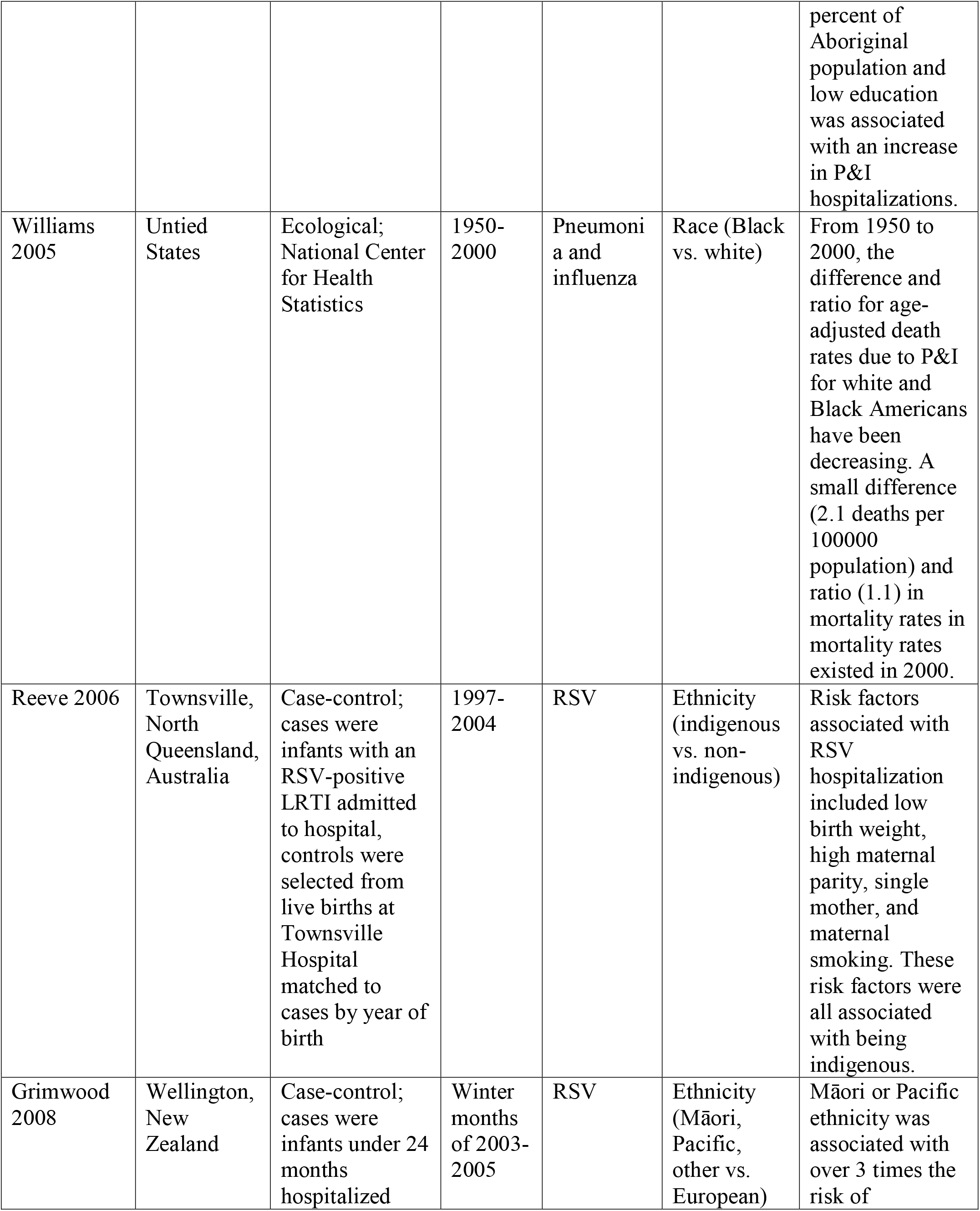
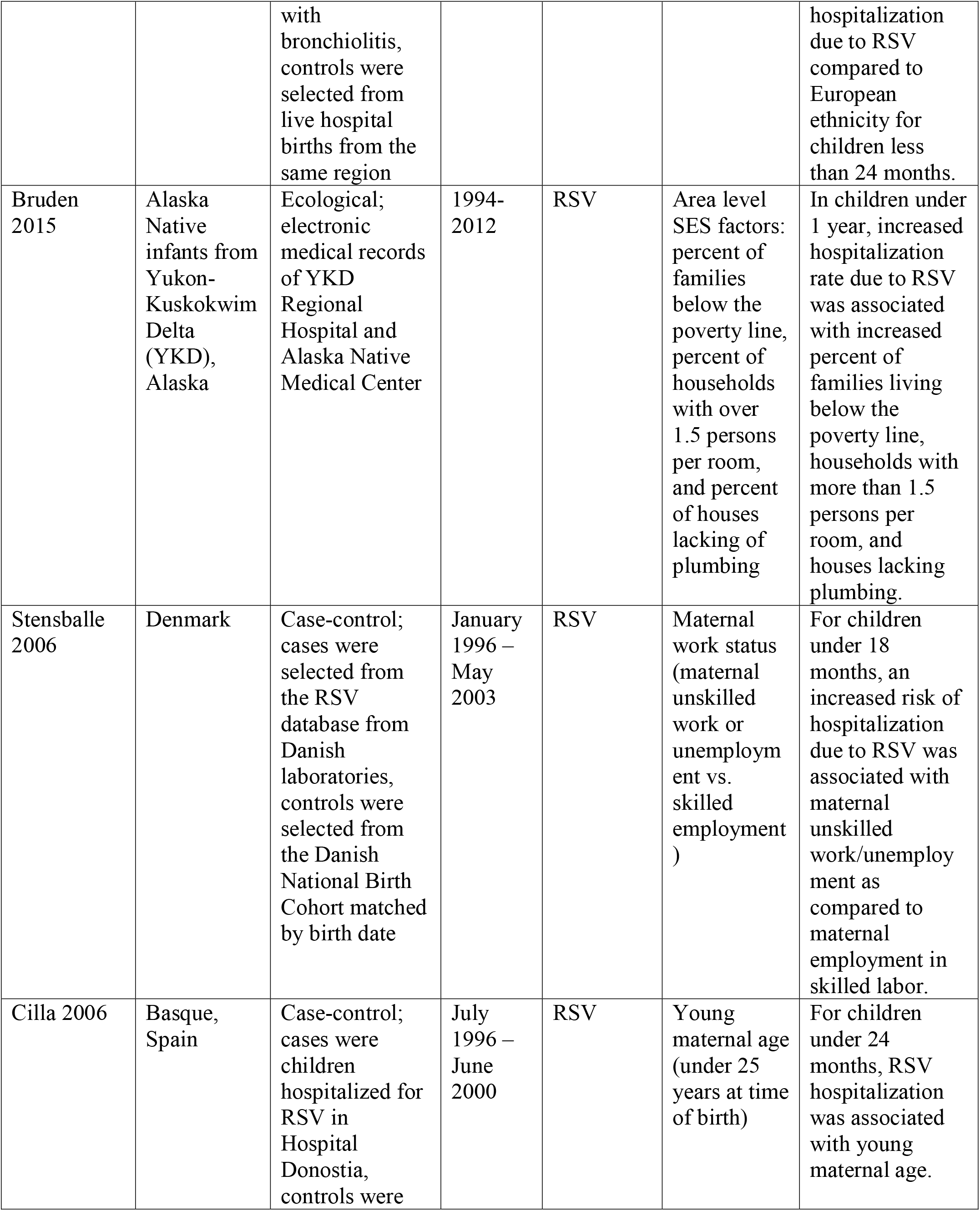
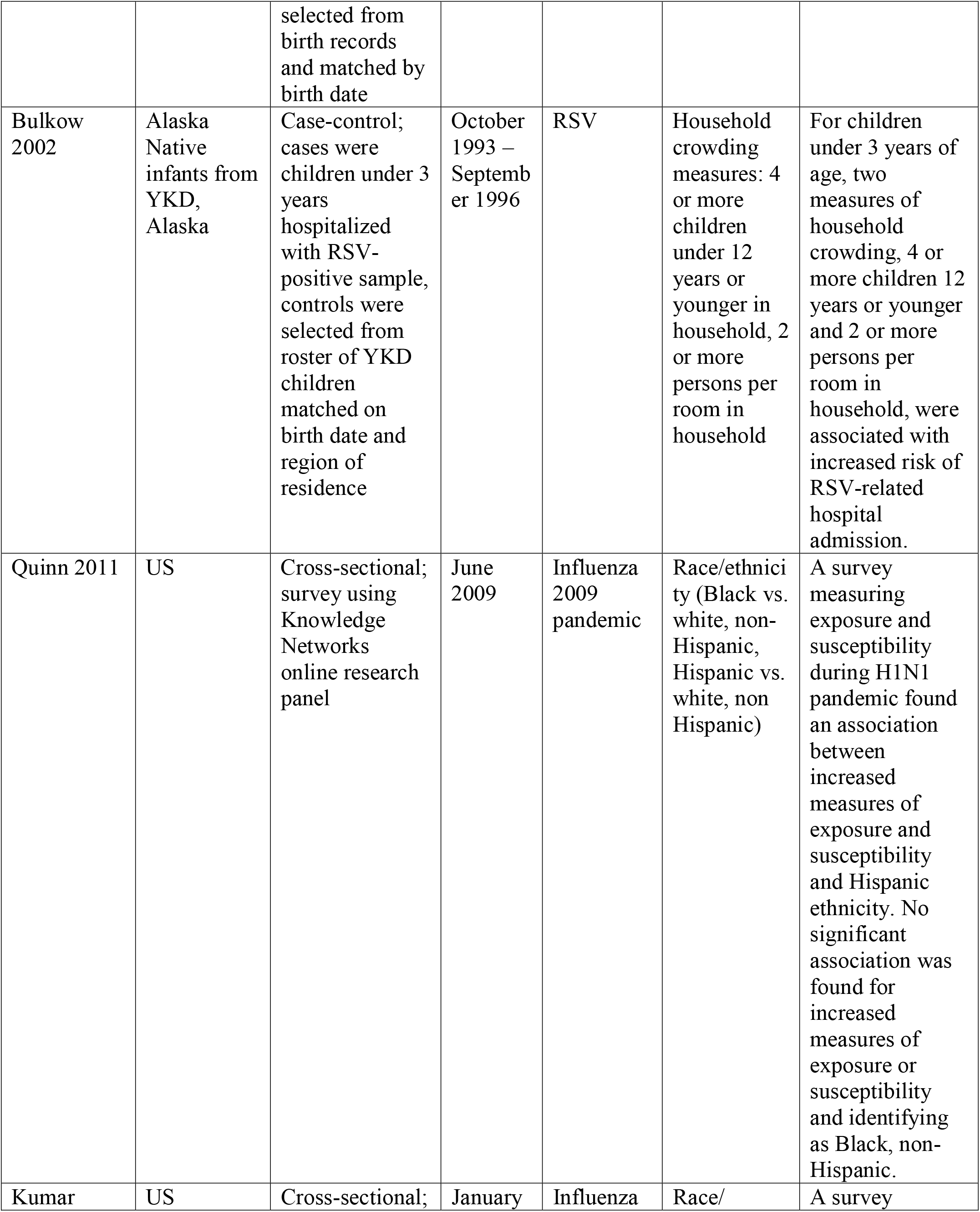
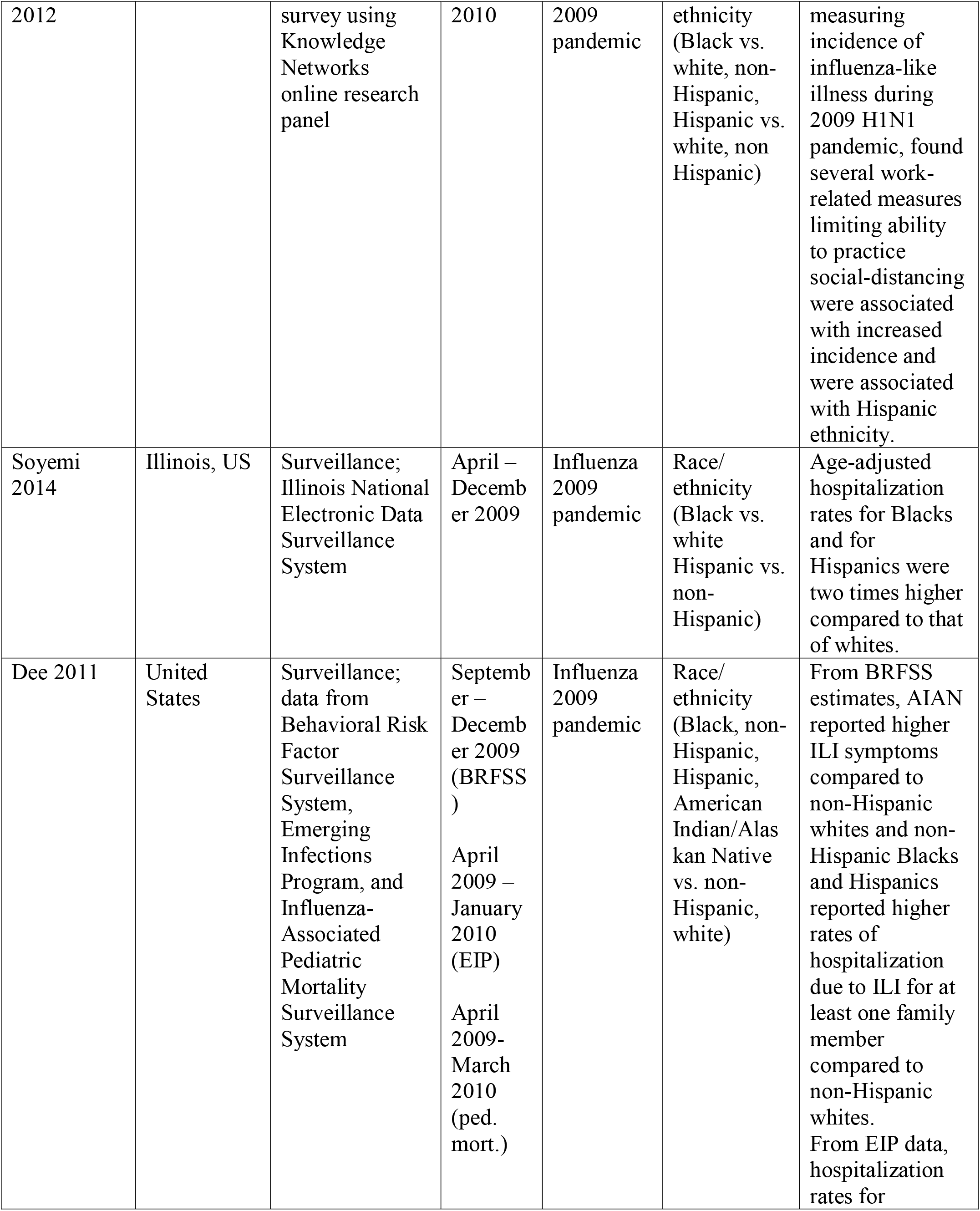
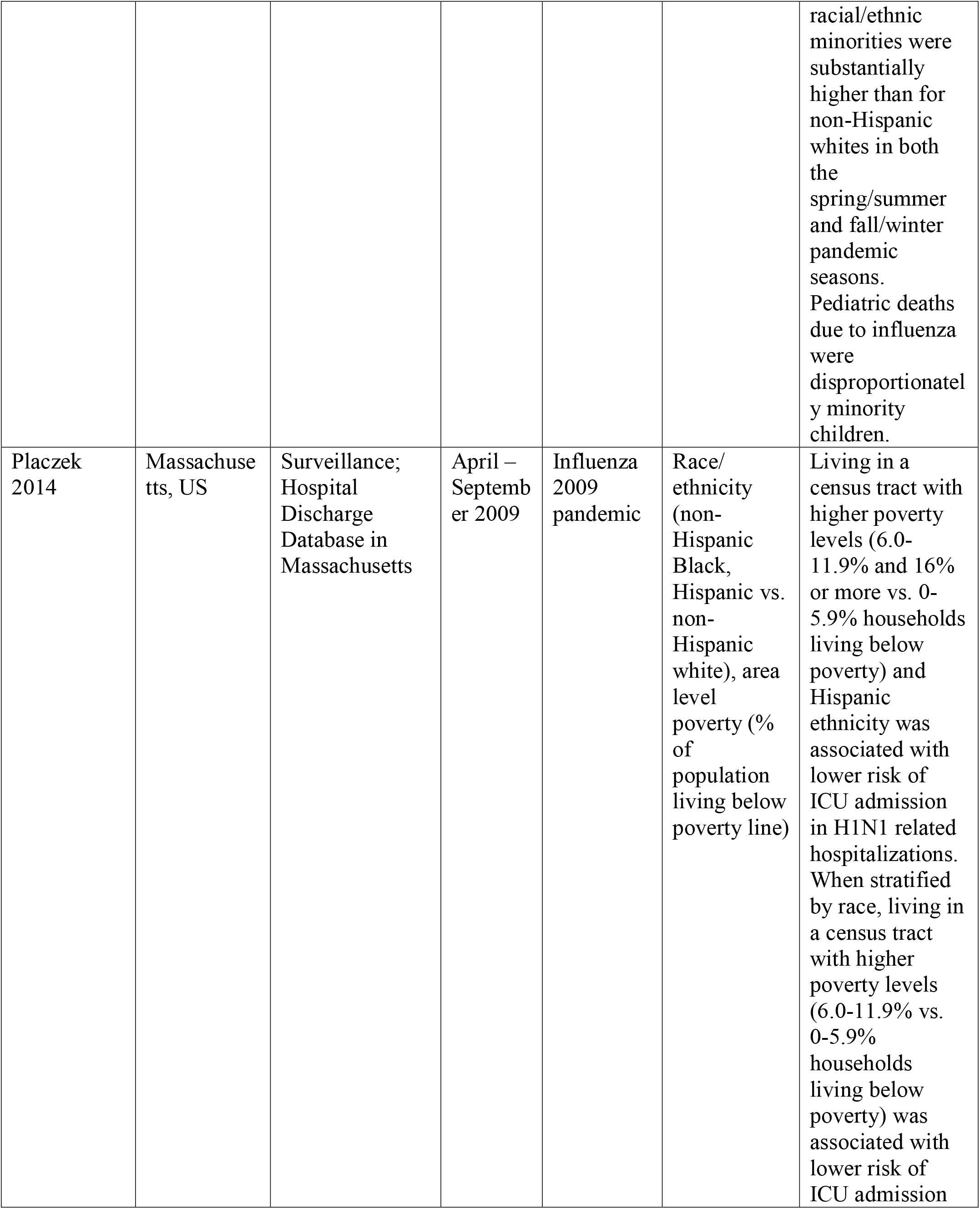
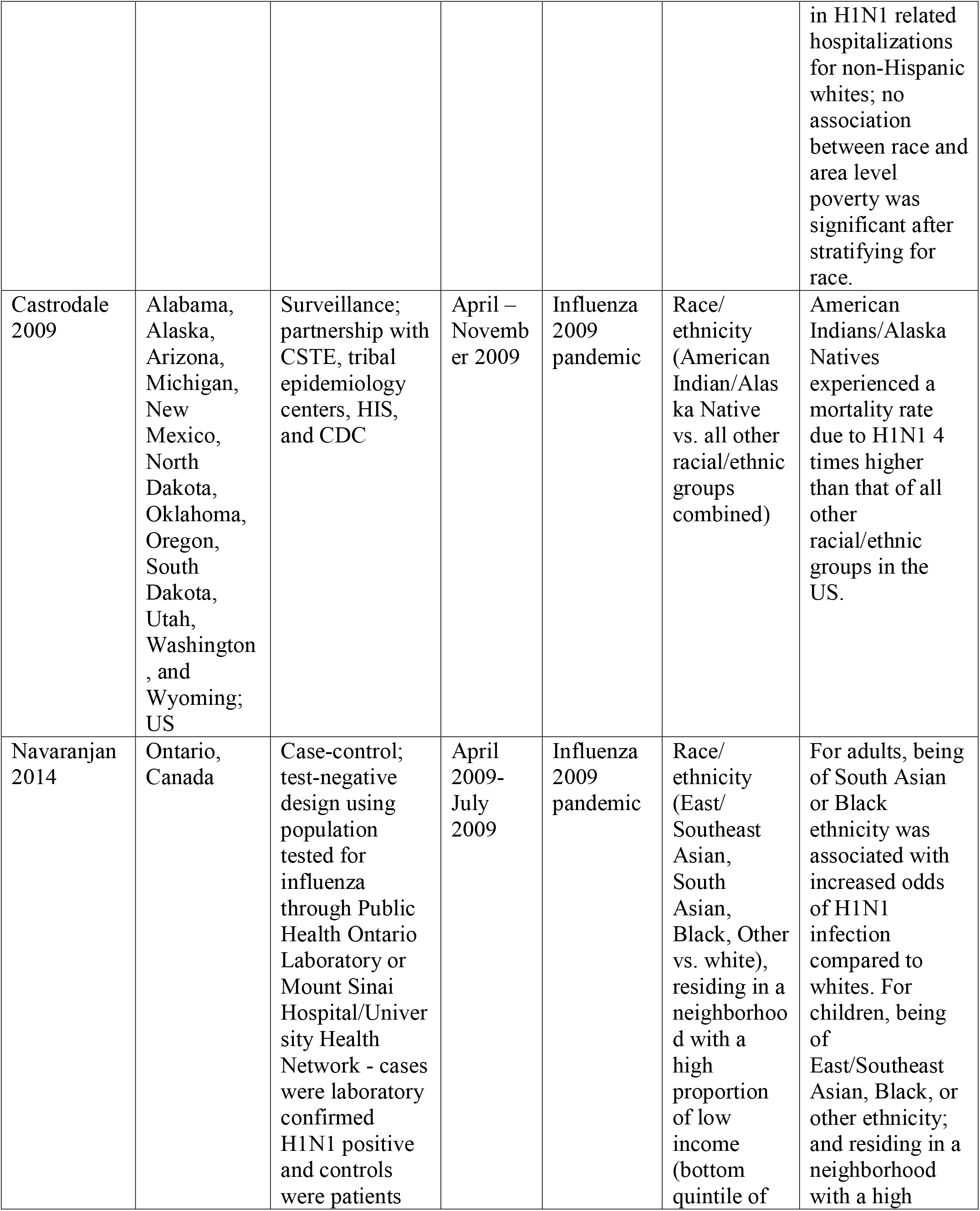
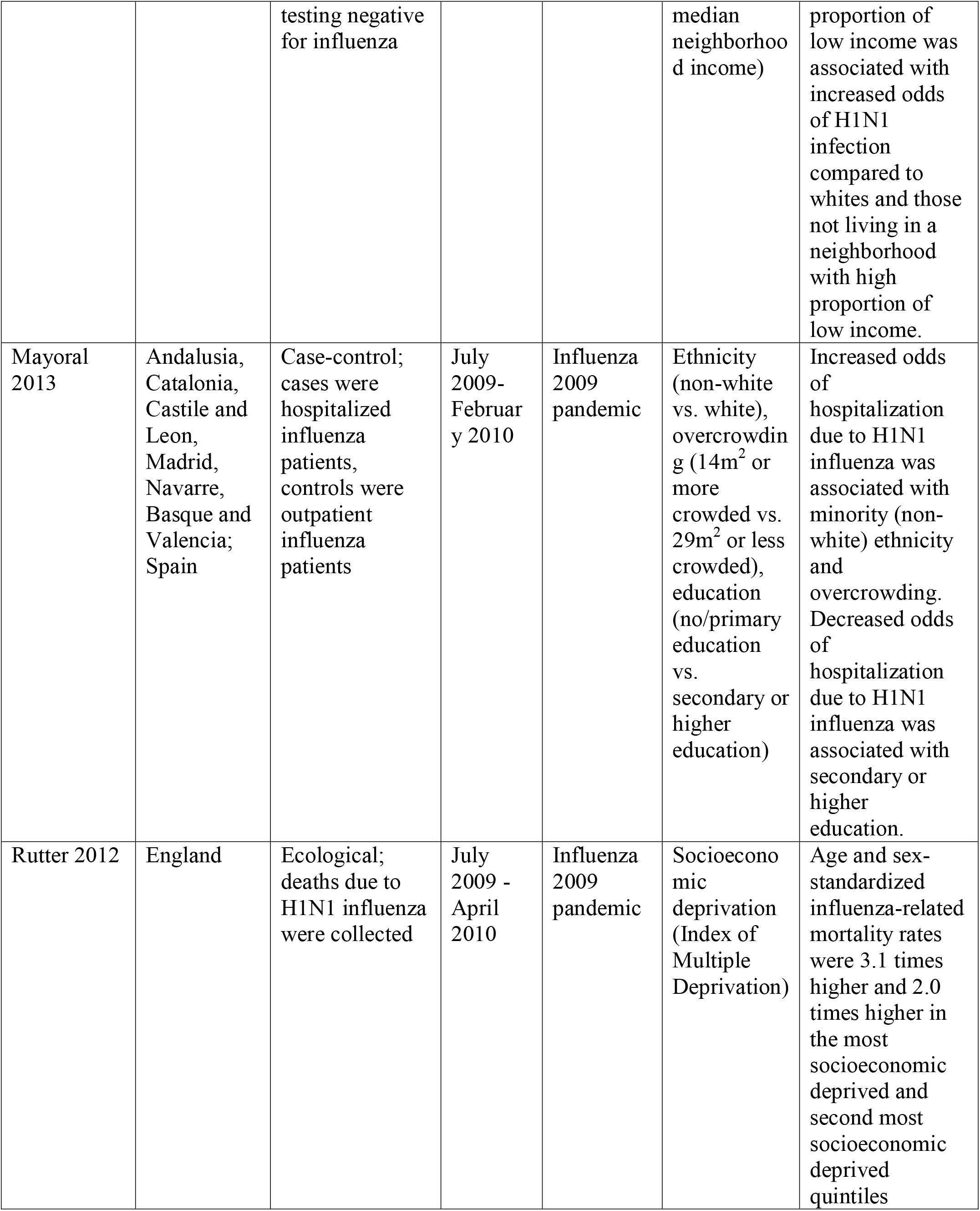
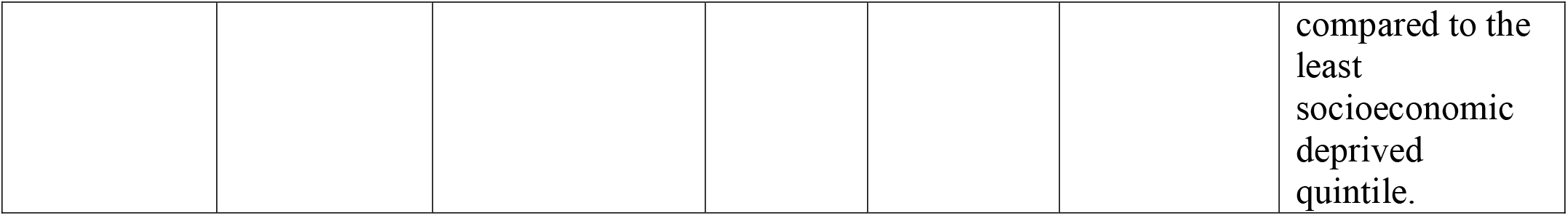
Study Key Findings.

## Data Availability

NA

## Supplementary Materials

### PubMed Search Terms

#### Search #1 (ARI/ILI/RSV Disparities)

((“delivery of health care”[MeSH Terms:noexp] OR “health behavior”[MeSH Terms] OR “health knowledge, attitudes, practice”[MeSH Terms] OR “health services accessibility”[MeSH Terms] OR “health services, indigenous”[MeSH Terms] OR “mass screening”[MeSH Terms] OR mass screening[TIAB] OR mass screenings[TIAB] OR health inequality[TIAB] OR health inequalities[TIAB] OR health inequities[TIAB] OR health inequity[TIAB] OR “health services needs and demand”[MeSH Terms] OR “patient acceptance of health care”[MeSH Terms] OR “patient selection”[MeSH Terms] OR “quality of health care”[MeSH Major Topic:noexp] OR “quality of life”[MeSH Terms] OR quality of life[TIAB] OR Culturally Competent Care[TIAB] OR “culturally competent care”[MeSH Terms] OR social disparities[TIAB] OR social disparity[TIAB] OR social inequities[TIAB] OR social inequity[TIAB] OR “socioeconomic factors”[MeSH Major Topic] OR socioeconomic factor[TIAB] OR socioeconomic factors[TIAB] OR “social determinants of health”[MeSH Terms]) AND (African American[TIAB] OR African Americans[TIAB] OR African ancestry[TIAB] OR “african continental ancestry group”[MeSH Terms] OR AIAN[TIAB] OR “american native continental ancestry group”[MeSH Terms] OR “asian continental ancestry group”[MeSH Terms] OR Asian[TIAB] OR Asians[TIAB] OR black[TIAB] OR blacks[TIAB] OR Caucasian[TIAB] OR Caucasians[TIAB] OR diverse population[TIAB] OR diverse populations[TIAB] OR environmental justice[TIAB] OR ethnic group[TIAB] OR “ethnic groups”[MeSH Terms] OR ethnic groups[TIAB] OR ethnic population[TIAB] OR ethnic populations[TIAB] OR ghetto[TIAB] OR ghettos[TIAB] OR Hispanic[TIAB] OR Hispanics[TIAB] OR Indian[TIAB] OR Indians[TIAB] OR Latino[TIAB] OR Latinos[TIAB] OR Latina[TIAB] OR Latinas[TIAB] OR “medically underserved area”[MeSH Terms] OR minority group[TIAB] OR “minority groups”[MeSH Terms] OR minority groups[TIAB] OR minority population[TIAB] OR minority populations[TIAB] OR Native American[TIAB] OR Native Americans[TIAB] OR “oceanic ancestry group”[MeSH Terms] OR pacific islander[TIAB] OR pacific islanders[TIAB] OR Native Hawaiian[TIAB] OR Native Hawaiians[TIAB] OR Alaska Natives[TIAB] OR people of color[TIAB] OR “poverty areas”[MeSH Terms] OR poverty area[TIAB] OR poverty areas[TIAB] OR “rural health”[MeSH Terms] OR rural health[TIAB] OR “rural health services”[MeSH Terms] OR “rural population”[MeSH Terms] OR rural population[TIAB] OR rural populations[TIAB] OR slum[TIAB] OR slums[TIAB] OR “urban health”[MeSH Terms] OR “urban health services”[MeSH Terms] OR “urban population”[MeSH Terms] OR urban population[TIAB] OR urban populations[TIAB] OR “vulnerable populations”[MeSH Terms] OR vulnerable population[TIAB] OR vulnerable populations[TIAB] OR white[TIAB] OR whites[TIAB]) OR (ethnic disparities[TIAB] OR ethnic disparity[TIAB] OR health care disparities[TIAB] OR health care disparity[TIAB] OR health disparities[TIAB] OR health disparity[TIAB] OR “health status disparities”[MeSH Terms] OR “healthcare disparities”[MeSH Terms] OR healthcare disparities[TIAB] OR healthcare disparity[TIAB] OR “minority health”[MeSH Terms] OR minority health[TIAB] OR sexual minorities[TIAB] OR racial disparities[TIAB] OR racial disparity[TIAB] OR racial equality[TIAB] OR racial equity[TIAB] OR racial inequities[TIAB] OR racial inequity[TIAB] OR “ageism”[MeSH Terms] OR “racism”[MeSH Terms] OR “apartheid”[MeSH Terms] OR “sexism”[MeSH Terms] OR “social discrimination”[MeSH Terms] OR “social segregation”[MeSH Terms] OR “social marginalization”[MeSH Terms]))

AND

(((((((Respiratory Syncytial Virus, Human[MeSH Terms]) OR RSV[Title/Abstract]) OR Respiratory Syncytial Virus[Title/Abstract]) OR Acute Respiratory Infection[Title/Abstract]) OR

ARI[Title/Abstract]) OR Influenza-like illness[Title/Abstract]) OR ILI[Title/Abstract]) OR influenza like illness[Title/Abstract])

### Search #2 (Influenza Disparities)

((“delivery of health care”[MeSH Terms:noexp] OR “health behavior”[MeSH Terms] OR “health knowledge, attitudes, practice”[MeSH Terms] OR “health services accessibility”[MeSH Terms] OR “health services, indigenous”[MeSH Terms] OR “mass screening”[MeSH Terms] OR mass screening[TIAB] OR mass screenings[TIAB] OR health inequality[TIAB] OR health inequalities[TIAB] OR health inequities[TIAB] OR health inequity[TIAB] OR “health services needs and demand”[MeSH Terms] OR “patient acceptance of health care”[MeSH Terms] OR “patient selection”[MeSH Terms] OR “quality of health care”[MeSH Major Topic:noexp] OR “quality of life”[MeSH Terms] OR quality of life[TIAB] OR Culturally Competent Care[TIAB] OR “culturally competent care”[MeSH Terms] OR social disparities[TIAB] OR social disparity[TIAB] OR social inequities[TIAB] OR social inequity[TIAB] OR “socioeconomic factors”[MeSH Major Topic] OR socioeconomic factor[TIAB] OR socioeconomic factors[TIAB] OR “social determinants of health”[MeSH Terms]) AND (African American[TIAB] OR African Americans[TIAB] OR African ancestry[TIAB] OR “african continental ancestry group”[MeSH Terms] OR AIAN[TIAB] OR “american native continental ancestry group”[MeSH Terms] OR “asian continental ancestry group”[MeSH Terms] OR Asian[TIAB] OR Asians[TIAB] OR black[TIAB] OR blacks[TIAB] OR Caucasian[TIAB] OR Caucasians[TIAB] OR diverse population[TIAB] OR diverse populations[TIAB] OR environmental justice[TIAB] OR ethnic group[TIAB] OR “ethnic groups”[MeSH Terms] OR ethnic groups[TIAB] OR ethnic population[TIAB] OR ethnic populations[TIAB] OR ghetto[TIAB] OR ghettos[TIAB] OR Hispanic[TIAB] OR Hispanics[TIAB] OR Indian[TIAB] OR Indians[TIAB] OR Latino[TIAB] OR Latinos[TIAB] OR Latina[TIAB] OR Latinas[TIAB] OR “medically underserved area”[MeSH Terms] OR minority group[TIAB] OR “minority groups”[MeSH Terms] OR minority groups[TIAB] OR minority population[TIAB] OR minority populations[TIAB] OR Native American[TIAB] OR Native Americans[TIAB] OR “oceanic ancestry group”[MeSH Terms] OR pacific islander[TIAB] OR pacific islanders[TIAB] OR Native Hawaiian[TIAB] OR Native Hawaiians[TIAB] OR Alaska Natives[TIAB] OR people of color[TIAB] OR “poverty areas”[MeSH Terms] OR poverty area[TIAB] OR poverty areas[TIAB] OR “rural health”[MeSH Terms] OR rural health[TIAB] OR “rural health services”[MeSH Terms] OR “rural population”[MeSH Terms] OR rural population[TIAB] OR rural populations[TIAB] OR slum[TIAB] OR slums[TIAB] OR “urban health”[MeSH Terms] OR “urban health services”[MeSH Terms] OR “urban population”[MeSH Terms] OR urban population[TIAB] OR urban populations[TIAB] OR “vulnerable populations”[MeSH Terms] OR vulnerable population[TIAB] OR vulnerable populations[TIAB] OR white[TIAB] OR whites[TIAB]) OR (ethnic disparities[TIAB] OR ethnic disparity[TIAB] OR health care disparities[TIAB] OR health care disparity[TIAB] OR health disparities[TIAB] OR health disparity[TIAB] OR “health status disparities”[MeSH Terms] OR “healthcare disparities”[MeSH Terms] OR healthcare disparities[TIAB] OR healthcare disparity[TIAB] OR “minority health”[MeSH Terms] OR minority health[TIAB] OR sexual minorities[TIAB] OR racial disparities[TIAB] OR racial disparity[TIAB] OR racial equality[TIAB] OR racial equity[TIAB] OR racial inequities[TIAB] OR racial inequity[TIAB] OR “ageism”[MeSH Terms] OR “racism”[MeSH Terms] OR “apartheid”[MeSH Terms] OR “sexism”[MeSH Terms] OR “social discrimination”[MeSH Terms] OR “social segregation”[MeSH Terms] OR “social marginalization”[MeSH Terms]))

AND

((((human influenza[MeSH Terms]) OR human influenzas[MeSH Terms]) OR human influenza[Title/Abstract]) OR human influenzas[Title/Abstract])

